# COVID-19: Tracking the Pandemic with A Simple Curve Approximation Tool (SCAT)

**DOI:** 10.1101/2020.04.06.20055467

**Authors:** Jane Courtney

**Affiliations:** Technological University Dublin, Ireland

**Keywords:** COVID-19, modelling, software, SARS-CoV-2, flattening the curve

## Abstract

In the current COVID-19 pandemic, much focus is put on ‘flattening the curve’. This epidemiological ‘curve’ refers to the cases versus time graph, which shows the rise of a disease to its peak before descending. The aim in a pandemic is to flatten this curve by reducing the peak and spreading out the timeline. However, the models used to predict this curve are often not clearly outlined, no model parameters are given, and models are not tested against real data. This lack of detail makes it difficult to recreate the curve. What is much needed is a simple tool for approximating the curve to allow ideas to be tested and comparisons made.

This work presents a Simple Curve Approximation Tool (SCAT) which can be used by anyone. This tool allows the user to approximate and draw the curve and allows testing of assumptions, trajectories and the wildly varying figures reported in the media. The mathematics behind SCAT is clearly outlined here but understanding of this is not required. SCAT is provided online as a downloadable MS Excel workbook with some sample cases shown. Throughout this work, the parameters used are specified so that all results can be easily reproduced.

Although not intended as a prediction tool, SCAT has achieved less than 0.5 % error in short-term forward prediction. It also shows a very significant improvement on the pandemic exponential approximations found throughout media reporting. As a comparison tool, it highlights obvious differences between COVID-19 and other diseases, such as influenza, and between countries at different stages of the pandemic (China, Italy and the UK are used here for demonstration purposes).

SCAT allows for quick approximation of the curve and creates meaningful comparisons and understandable visualisations for COVID-19 and other diseases.

## Introduction

The coronavirus pandemic 2019 (COVID-19) has now spread to almost all countries in the world and has infected over 1.2 million people to date and killed almost 70,000.^1^ Despite this, many governments have been slow to react to the crisis. Figure 1 shows the numbers of cases per million before government interventions.

**Figure 1:**
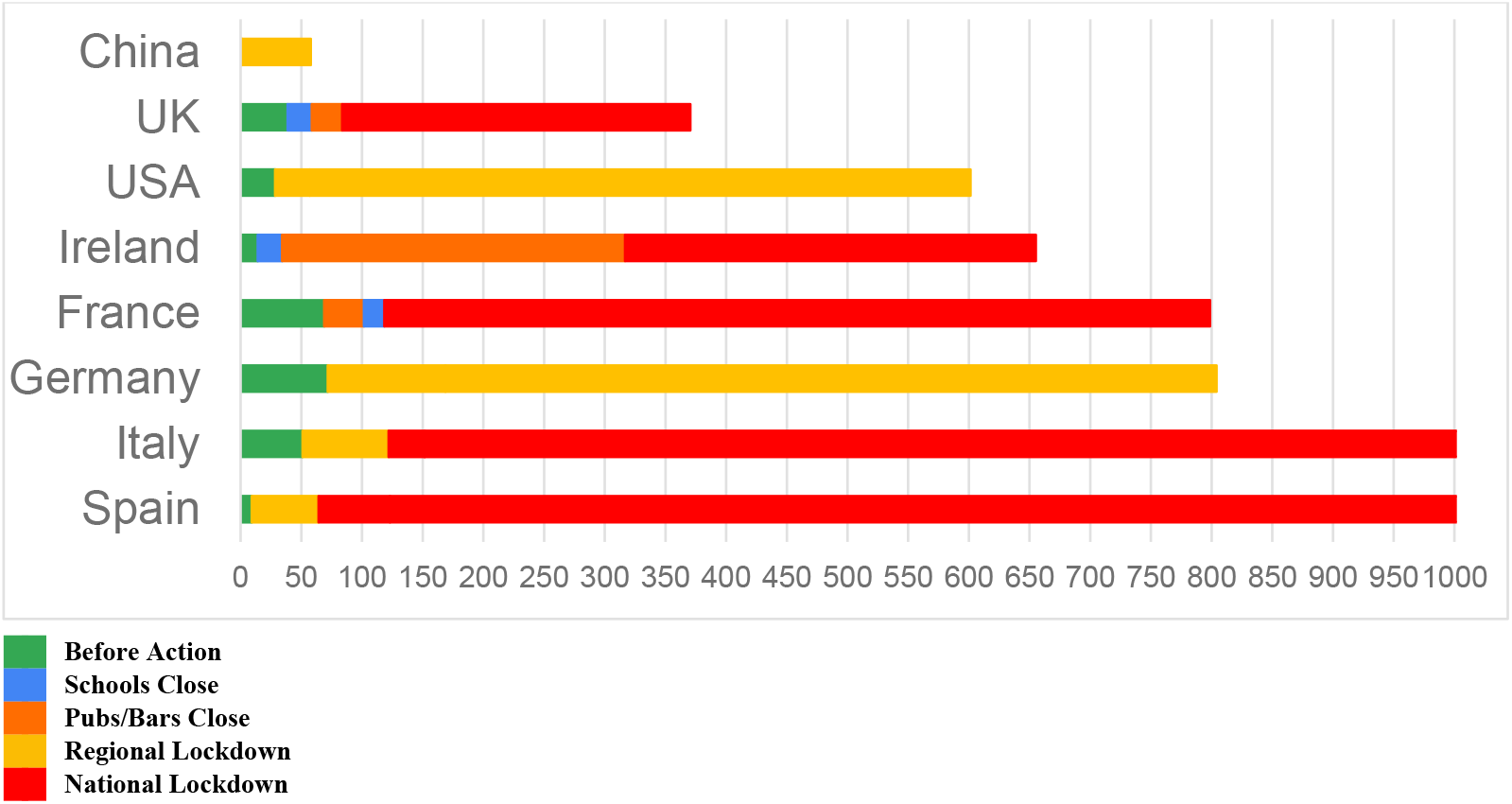
*Cases per million population before government intervention. ‘Before Action’ may include significant contact tracing and testing approaches. This graph shows just closures and lockdowns.*

Those governments that have been slower to react have cited varied sources in explaining their reasons for inaction. For example, in the UK, the much cited Imperial College Report^2^ has been used to guide national policy. Although the epidemiological curve is presented in this report, no details are given as to how it was produced, making it impossible to recreate. Nonetheless, the model is based on previous work^3^ and has considerable merit. It uses population density, household data, school data, workplace data as well as commuting and travel data to ultimately construct the curve. This is certainly useful when modelling an epidemic in a country but is quite difficult to extend to a pandemic or to recreate for different diseases or populations.

Traditional epidemiological models use variations on the susceptible-infected-recovered (SIR) model.^4^ This model approximates the basic reproduction number, *R*_0_, the expected number of new infections arising from each infection. To estimate this, the time delay between infection and diagnosis and between infection and infectiousness need to be known or assumed. The difficulty with this model is twofold. Firstly, it requires a good understanding of epidemiology to produce the elusive ‘curve’, and secondly, it can be misled by incorrect assumptions of initial conditions, and any errors will quickly propagate through the model.^5^

As a result, many authors and reporters have reverted to much simpler models, typically exponential increases.^6^ However, these models break down as the curve begins to flatten,^7^ either due to interventions or as a natural progression of the disease. The assumption that the infection rate will only increase is also leading to unrealistic figures reported in the media.^8^ Between inactive governments and irresponsible sources underplaying the numbers and describing the virus as ‘just the flu’^9^ and the exponential models describing an apocalyptic scenario,^10^ it is difficult to eke the truth out from the reporting.

While epidemiologists seek to understand the behaviour of the disease^11^, for many applications outside this field, a simple approximate curve will suffice. Mathematicians, engineers and others may recognise the curve as having a somewhat Gaussian shape^12^. Although this is not entirely true, a Gaussian curve does fit the data with reasonable accuracy, particularly in the rise phase. It also has the advantage of being defined by clearly understandable and adjustable parameters. This type of Gaussian model is in fact often used in epidemiological approximations,^13^ and has been used to approximate other pandemics such as AIDS.^14^ It should be noted that this model is only approximate, making it useful for visualisation but not for long-term predictions as was done in the AIDS study.^15^

However, as presented in the results here, this model fits well to a fast-rising pandemic like COVID-19 and serves as a useful visualisation tool for approximating the curve.

## Methods

The Simple Curve Approximation Tool (SCAT) presented here extrapolates the curve based on currently available data. This allows the pandemic to be tracked in a visually understandable way, allowing comparisons to be made with other diseases and across countries at different stages of the pandemic.

Short-term prediction is accurate with less than 0.5% error when modelling the UK curve based on data taken 20 days earlier. It is not intended that this would be used for long-term prediction, as it is certainly not as sophisticated as complex epidemiological models.^16–18^

Any user can modify the curve based on new data as SCAT is available freely online as a downloadable MS Excel workbook with some sample data provided for guidance. Figure 2 shows examples of curves generated by SCAT, showing the original and flattened curve now so very familiar in COVID-19 reporting.

**Figure 2:**
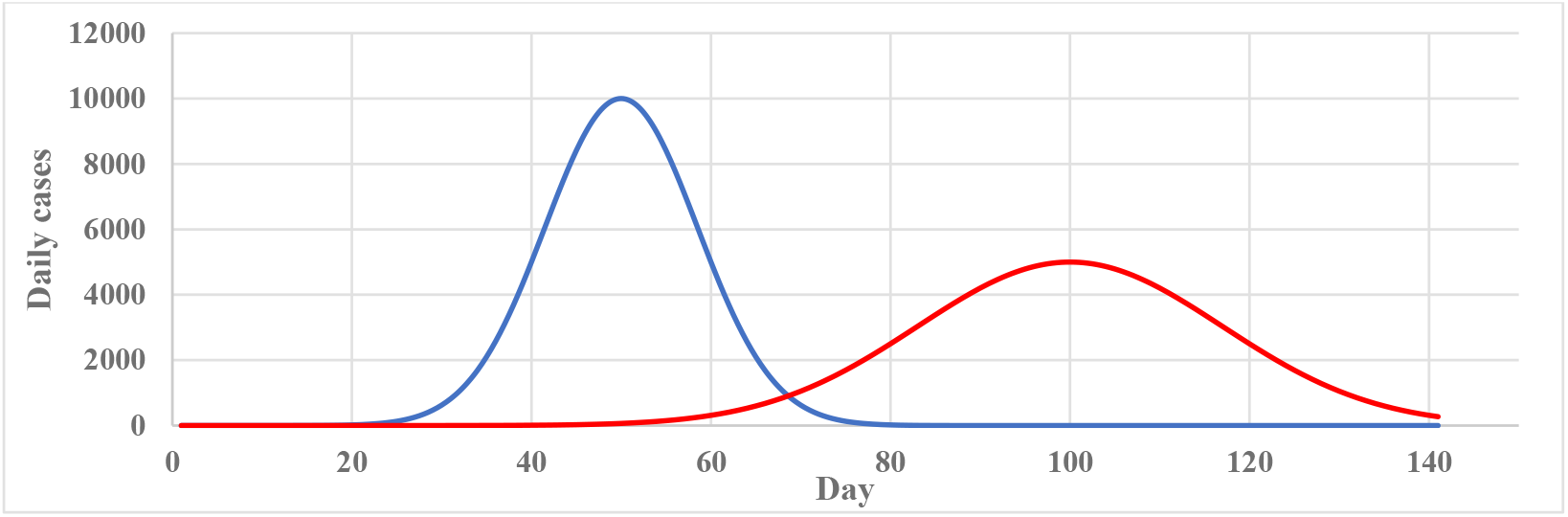
*Variations on the curve generated by SCAT.*

SCAT can be used for verifying media reported figures, or simply for drawing the curve for publications. It should be noted that no causation is attached to any findings nor are any hypothetical scenarios investigated. The results presented here simply extrapolate from what is, based on currently available data.

### Defining the Curve

While the equation of the curve may not be familiar, its parameters are easily understood. The curve can be described by the following Gaussian function:

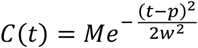

*C*(*t*) represents the number of cases per day

*t* is the time (day number since data recording began)

*M* is the maximum number of cases recorded in any one day

*p* is the peak day, i.e. the day on which the maximum *M* occurs

*w* is a parameter related to the width of the curve

The parameter *w* is proportional to the Full Width at Half Maximum (FWHM), i.e. the width of the curve at half *M*, and is calculated as:

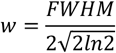

These parameters can be seen on the curve in Figure 3. Here, a maximum day rate of 6,000 is shown at day 40 with a curve width (FWHM) of 20. In SCAT, these figures can be easily modified to give different curves. In all results in this work, parameters are shown so they can be easily recreated.

**Figure 3:**
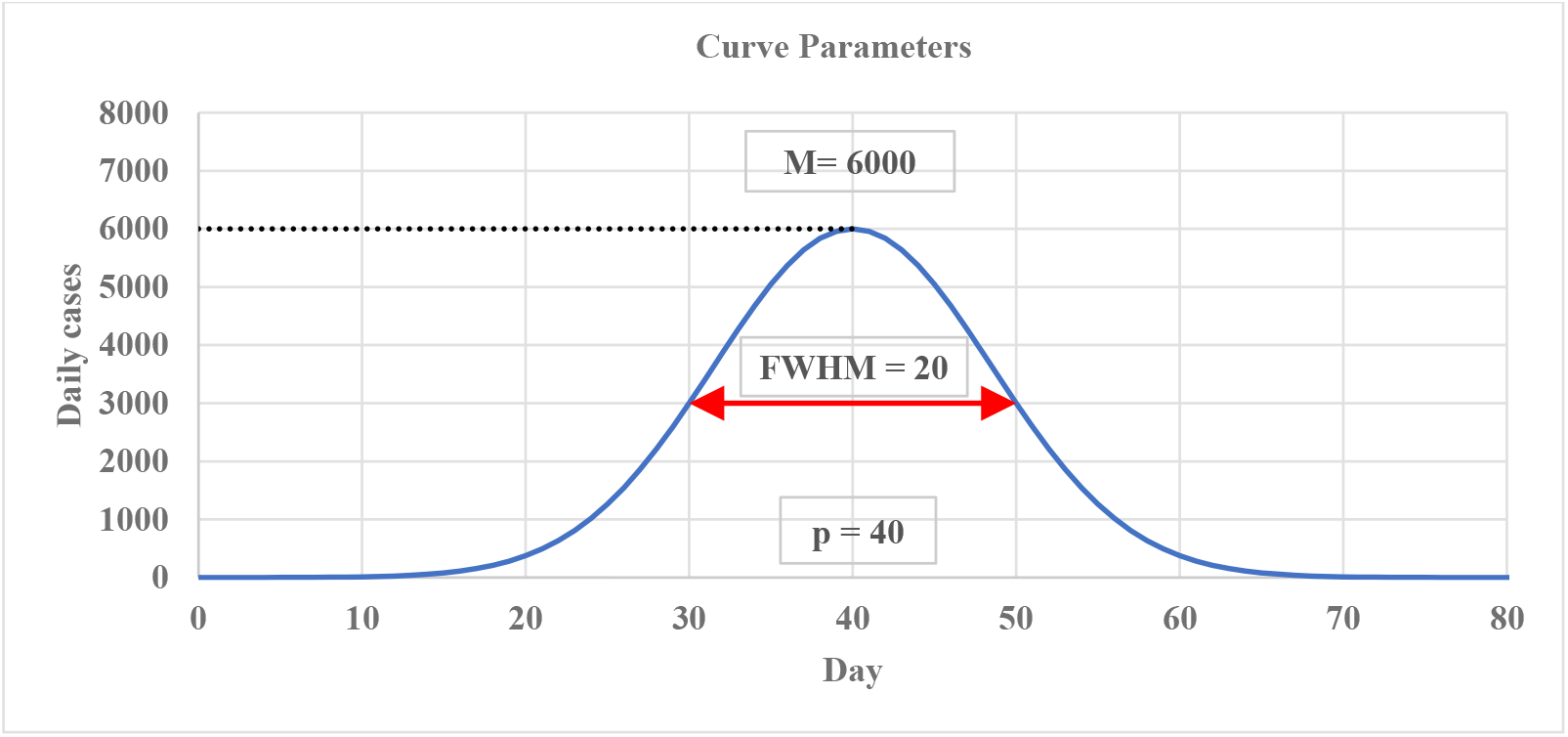
*The curve showing its parameters M* = 6000, *p* = 40, *FWHM* = 20

## Results

All real data used for testing and performance analysis was gathered from WHO statistics,^19^ using Day 1 = 1 January 2020 up to Day 92 = 1 April 2020. Results are presented here for some of the many applications of SCAT. These include comparisons across countries at different stages of the pandemic and with different policies in place, testing the flattening of the curve, approximating when healthcare system capacity will be reached, comparison with other diseases and short-term prediction.

To test SCAT, Chinese data was used, as China has seen a peak of cases and returned to pre-pandemic levels. This is used simply to validate the proposed curve. Due to a shift in definition of diagnosis on 12 February 2020, an outlier is observed on 13 February 2020 which causes the data to deviate from the curve. Otherwise, it fits the expected Gaussian shape. Figure 4 shows the results.

**Figure 4:**
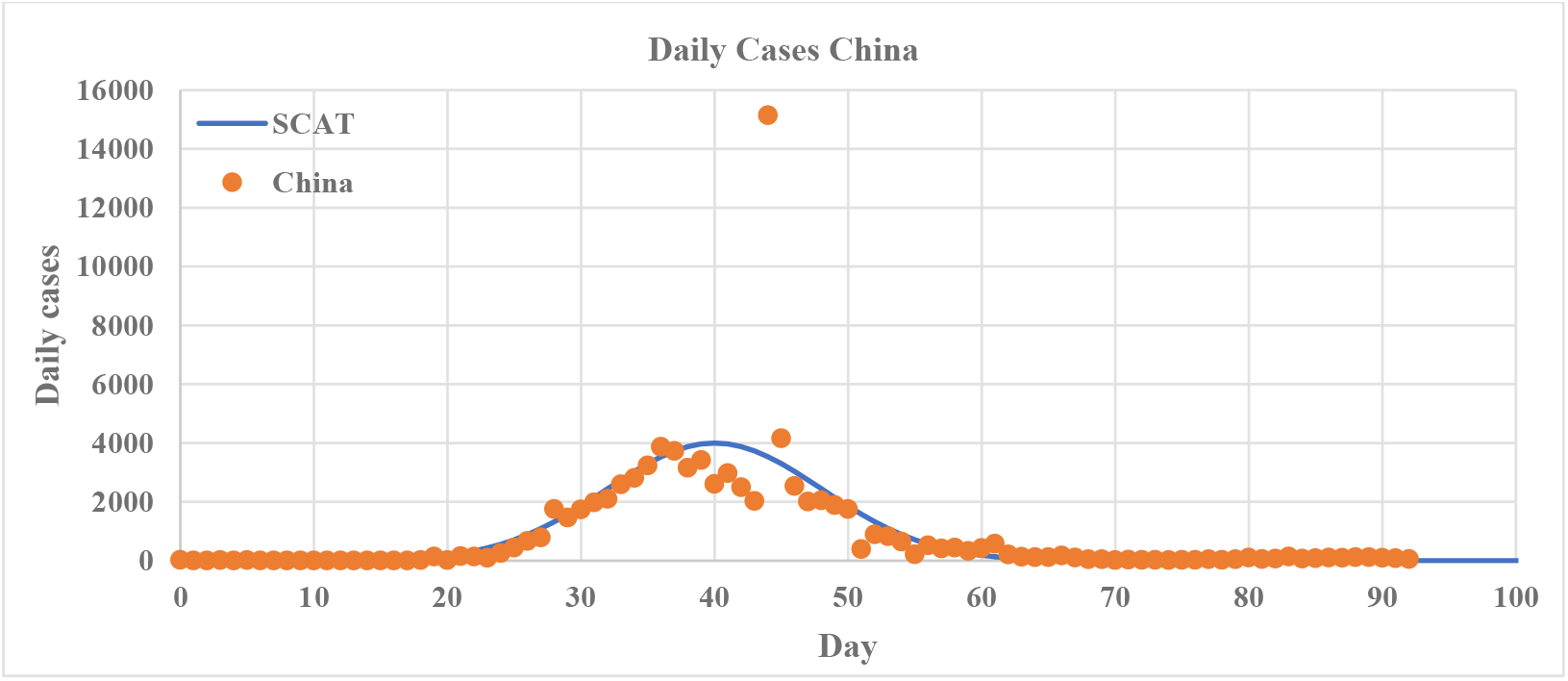
*SCAT with parameters M* = 4000, *p* = 40, *FWHM* = 19 *applied to Chinese data. Data is recorded from Day 1 = 1 January 2020 so the peak occurred at Day 40 = 8 February 2020 (Note: an outlier on 13 February 2020, due to a redefinition of diagnosis, causes the data to deviate from the curve).*

Although SCAT is not particularly exact throughout (this is not its intention), it does a reasonable approximation of the rise phase. This is significant as it shows it can be used for approximating the pandemic in other countries where the number of cases is still rising. Despite deviation from the data in parts, it also shows accuracy when predicting the total number of cases (See Figure 5).

**Figure 5:**
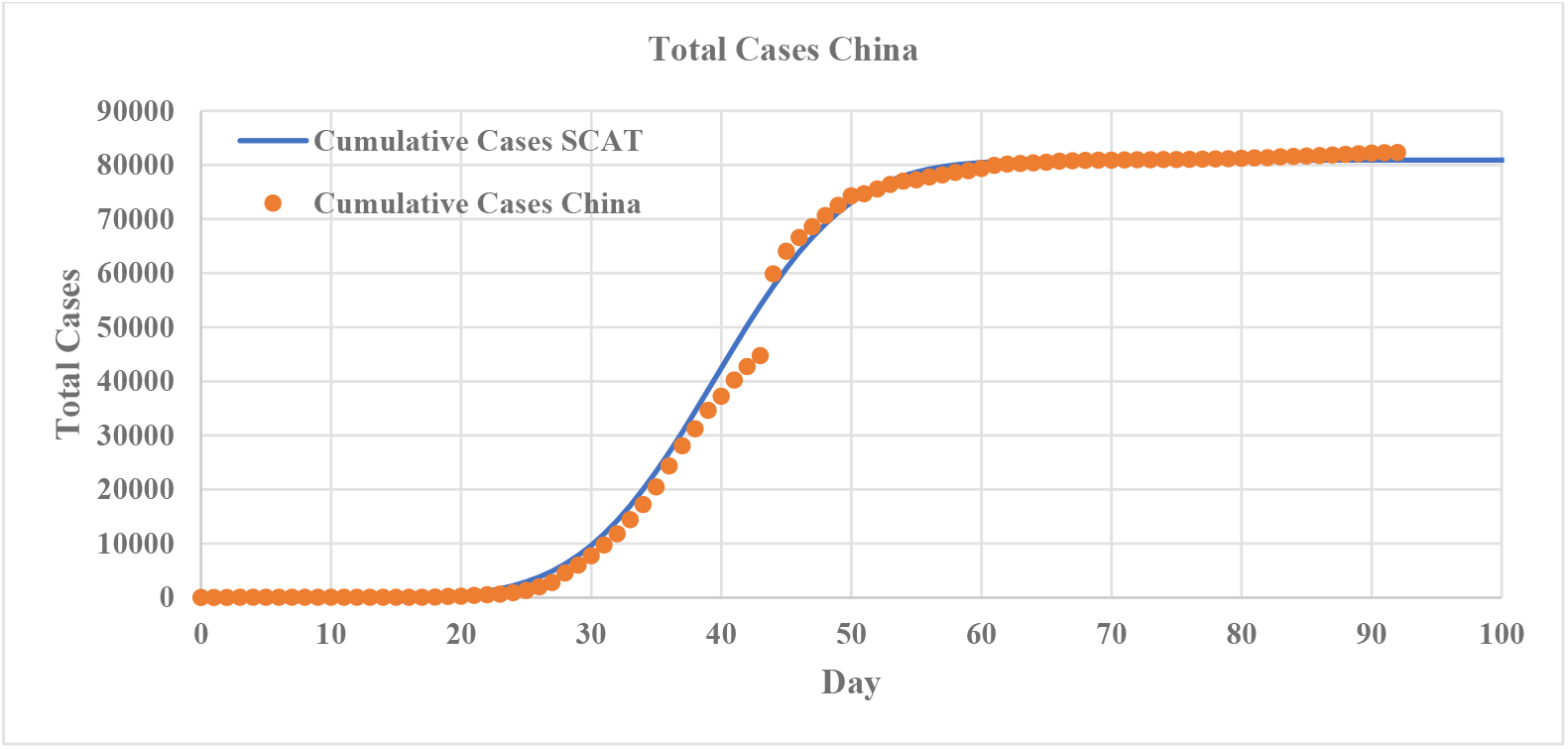
*SCAT applied to China* (*M* = 4000, *p* = 40, *FWHM* = 19) *showing cumulative results.*

SCAT was also applied to Italian cases, with parameters adjusted to fit the data. The width (*FWHM*) of the Chinese curve is likely to be narrower than countries affected later. As the virus spread to Italy after China, the peak day (*p*) will be later. Finally, the maximum number of cases recorded in any one day (*M*) is adjusted until the curve approximately fits the data. The Results are shown in Figure 6.

**Figure 6:**
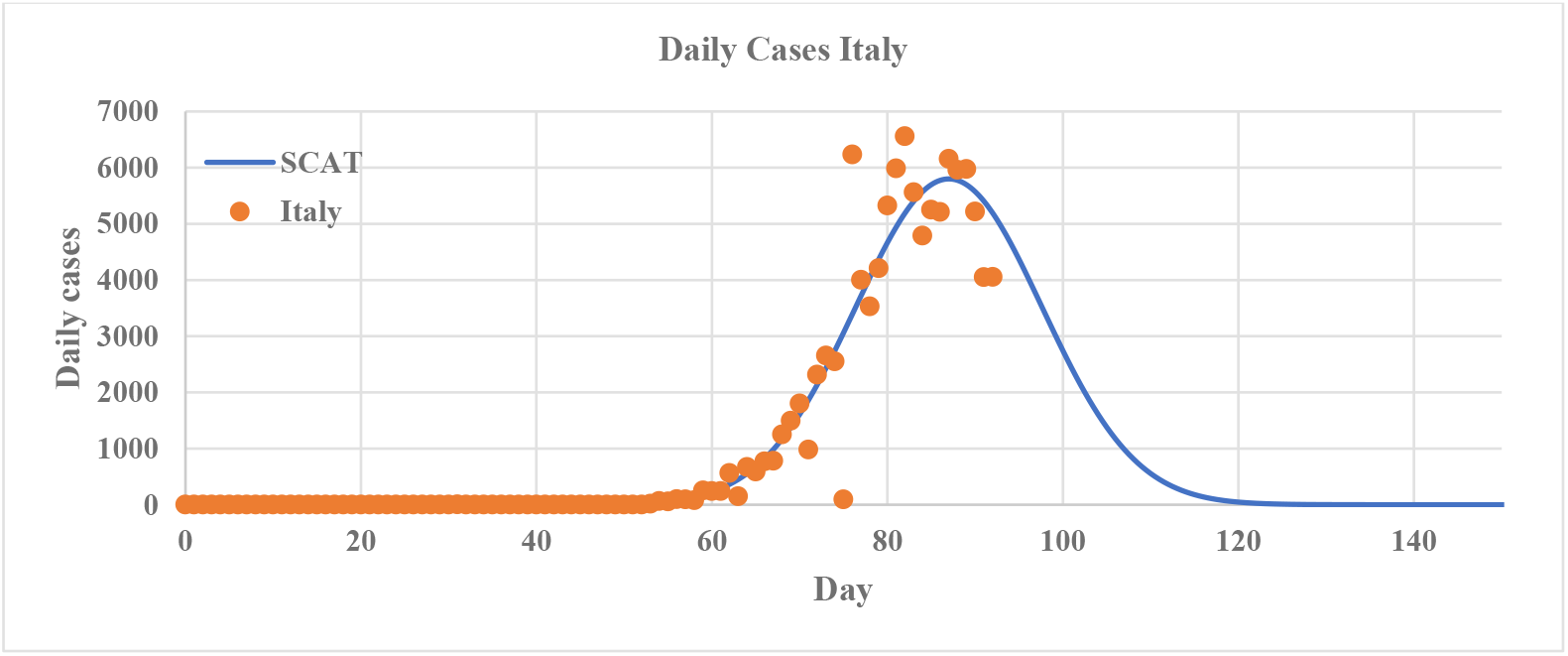
*SCAT applied to Italy* (*M* = 5800, *p* = 87, *FWHM* = 25).

Besides the anomaly observed in China due to a redefinition of diagnosis, there are other issues with tracking cases. Some countries have rolled out significant testing measures which means that a vast number of cases are captured. Others have more recently had to curtail testing. There are also dates on which little or no data was recorded. This is more likely due to a glitch in the data gathering and reporting than a lack of cases on those days, as the subsequent day often captures this lost data. This means that the level of testing, and subsequently the data is not consistent throughout the dataset.

However, SCAT can also be used in tracking deaths, which tend to be easier to confirm. It should be noted, however, that comorbidity is not considered in the data reporting of COVID-19. This is significant, as the demographic with the highest COVID-19 mortality rates (those over 80 years of age) also naturally has the highest morality rates due to other causes.

In general, it is observed that in countries in the early stages, where testing has not been curtailed, the pandemic can be tracked best using cases, whereas deaths allow for better tracking in countries at a later stage, when testing has not remained consistent. Figure 7 shows this to be the case for Italy, with daily deaths adhering more closely to the curve.

**Figure 7:**
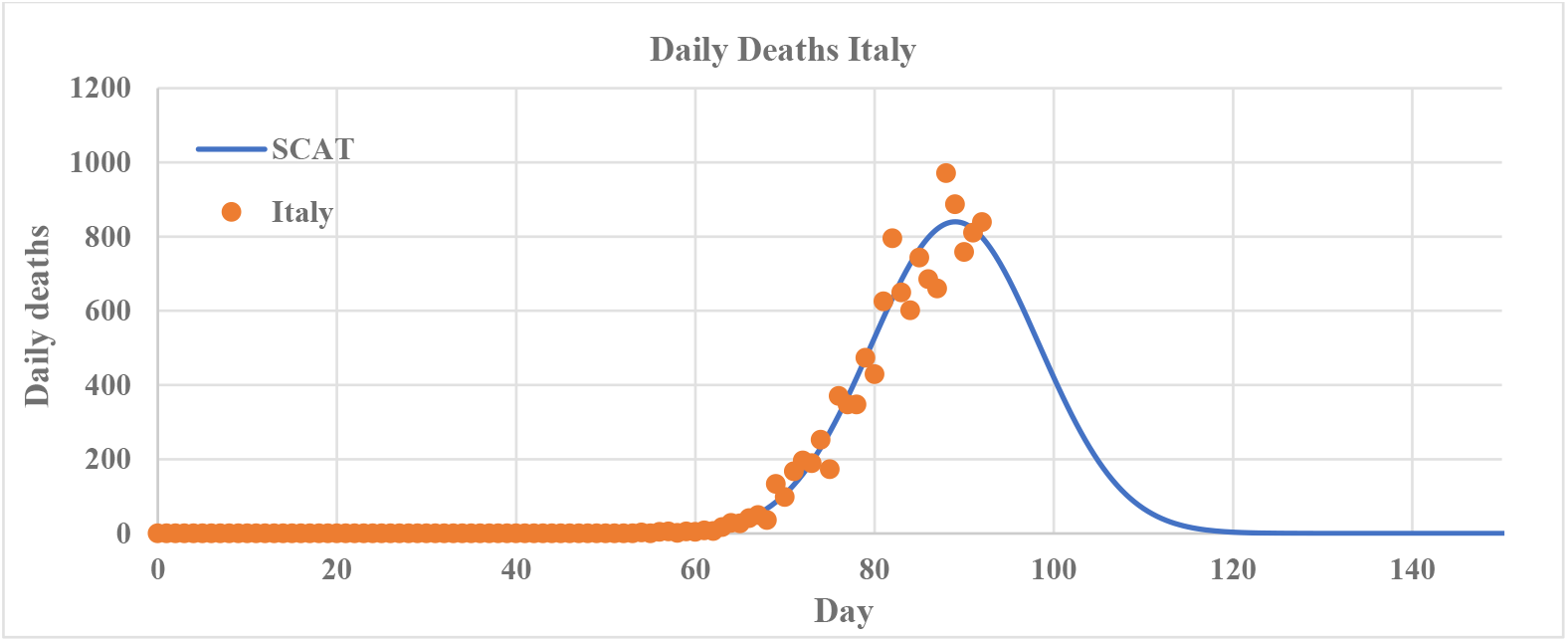
*SCAT applied to Italian deaths* (*M* = 840, *p* = 89, *FWHM* = 22).

### Short-term Prediction

Although not intended for long-term prediction, SCAT has shown surprising accuracy in short-term forecasting for countries that have not attempted to flatten the curve. For example, the UK put no social distancing measures or government mandates in place until 18 March 2020 (day 77).^20^ SCAT was applied the UK data on 11 March 2020 (day 71), with parameters *M* = 8100, *p* = 108, *FWHM* = 29 (See Figure 8). The curve was then revisited twenty days later, on 31 March 2020 (day 91), and less than 0.5 % error was observed. Results are shown in Table 1.

**Table 1:** *SCAT applied to UK data on 11 March 2020 and compared on 31 March 2020, showing less than 0.5% error.*

| Date | Recorded Total Cases | SCAT Prediction | Error (%) |
| --- | --- | --- | --- |
| 11 March 2020 | 373 | 374.1855 | 0.318 |
| 31 March 2020 | 22,141 | 22,249.154 | 0.488 |

**Figure 8:**
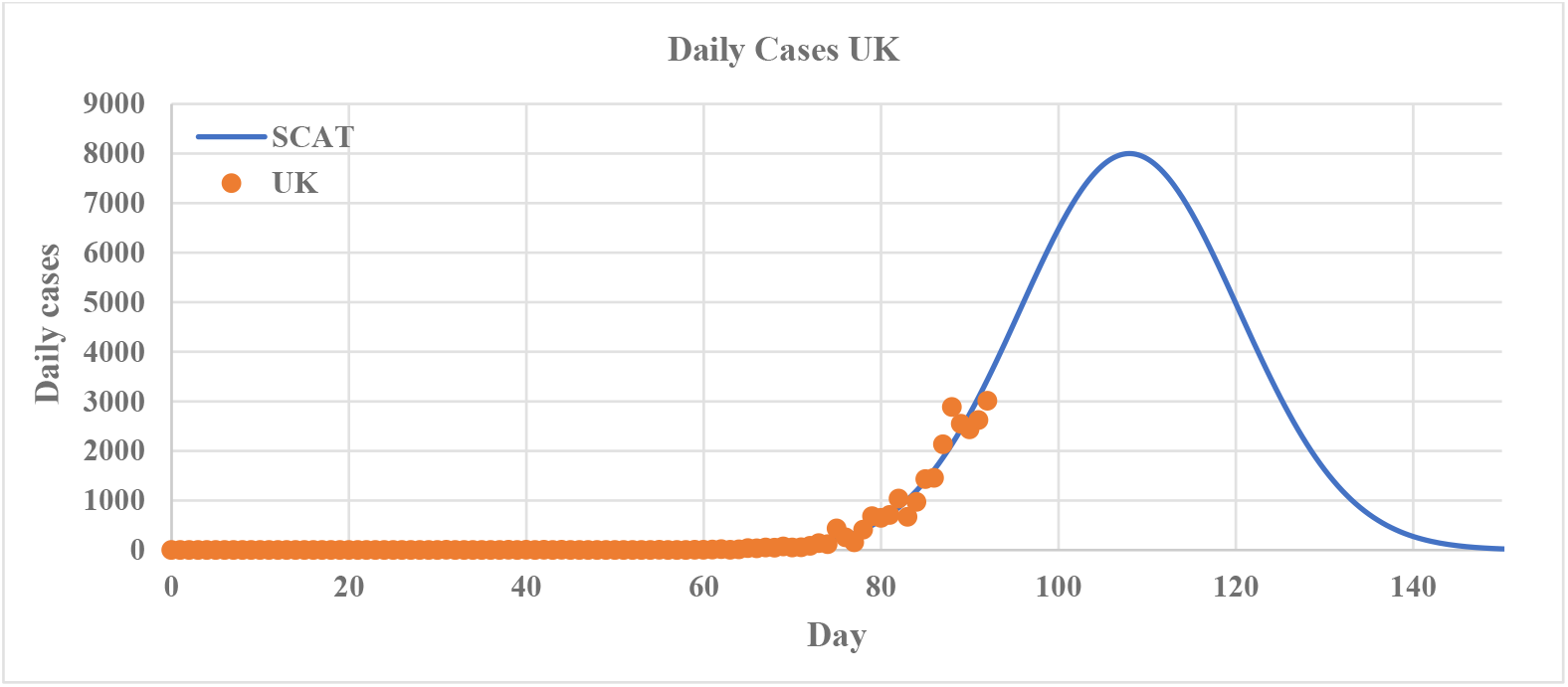
*SCAT applied to the UK* (*M* = 8000, *p* = 108, *FWHM* = 29).

It should be reiterated, however, that SCAT should not be used for long-term prediction as diseases are rarely well behaved over time. This has been done erroneously with the AIDS pandemic, producing misleading results.^15^ It is also hoped that the measures now in place will have a positive effect, so the recorded data should start deviating from the curve.

### Comparison Between Countries and Curve Flattening

The question on everyone’s minds is ‘Are we flattening the curve?’^21^

To test this, SCAT is used to compare countries that were affected earlier by the pandemic with countries affected later.^22^ In Europe, Italy has been taken as a starting point for comparison as it was one of the earliest affected countries. Many countries are on a similar trajectory but at an earlier stage of the pandemic and any deviations from that trend may suggest a flattening of the curve or otherwise.

As mentioned previously, the UK was relatively slow to react to the crisis, implementing its first social distancing measures on 18 March 2020 (day 77) when 2,630 cases were already recorded in the country (approximately 39 cases per million). When comparing the UK data to the Italian curve a similar trajectory is observed (See Figure 9).

**Figure 9:**
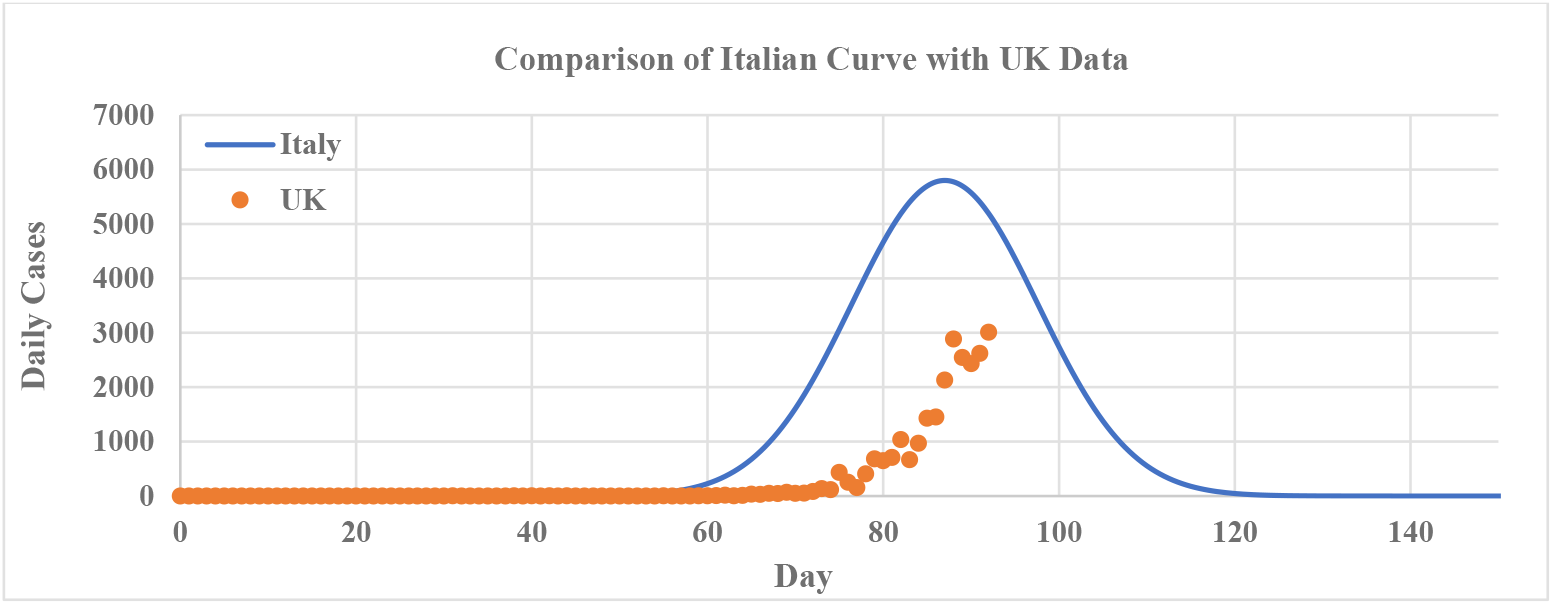
*Comparisons of the Italian curve* (*M* = 5800, *p* = 87, *FWHM* = 25) *with the UK data. The Italian curve is not a good fit but shows a similar trajectory to the UK.*

By viewing curves on the same timeline, the curves for different countries can be easily compared (see Figure 10).

**Figure 10:**
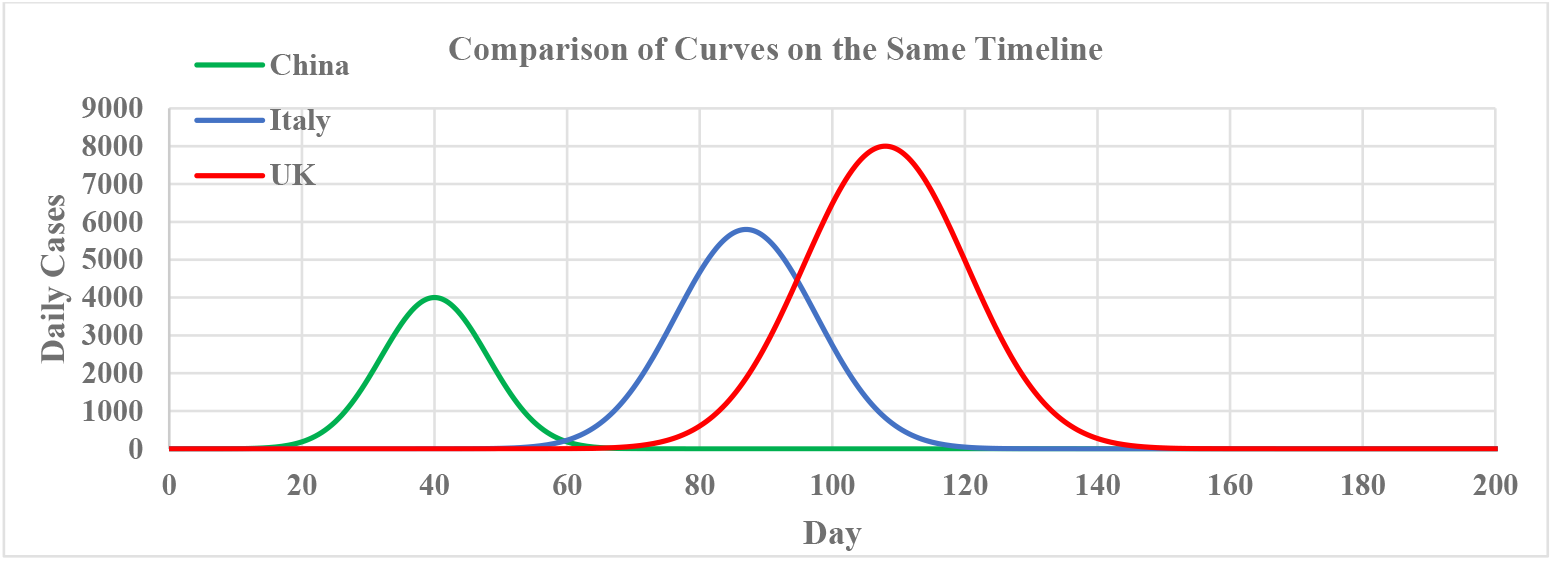
*Comparisons of the curves for China* (*M* = 4000, *p* = 40, *FWHM* = 19), *Italy* (*M* = 5800, *p* = 87, *FWHM* = 25) *and the UK* (*M* = 8000, *p* = 108, *FWHM* = 29).

### Hospital Capacity

The real danger of COVID-19 is not the ultimate infection or death rate but the likelihood that healthcare systems will become overwhelmed. This is what has been observed in Italy, where the pandemic has reached crisis levels. To identify the point at which this overwhelming occurs, a line is often drawn on the curve at healthcare system capacity.

However, it can be difficult to approximate this capacity for several reasons. Firstly, the number of critical care beds available is dynamic depending on occupancy rates. Secondly, the number of active COVID-19 cases is also dynamic as patients either die or recover. Also, most cases of COVID-19 are mild and do not require hospitalisation so using the daily cases curve is not necessarily helpful. Finally, the national figures for overall healthcare system capacity do not reflect the geographical spread – while the hospitals in Lombardy were overwhelmed, it would not be feasible to send a critical care patient to Sicily or Puglia to be treated if that’s where critical care beds were available.

Since the purpose of SCAT is to draw the curve, a crude attempt is made here to approximate healthcare system capacity and draw a capacity line on the curve. In Italy, approximately 10% of infected patients were admitted to intensive care units daily during early to mid-March.^23^ Before the construction of COVID-19 hospitals, the capacity of Italy’s available critical care was around 5,200 beds. Using these approximations suggests that Italy’s capacity for COVID-19 cases would have been 52,000. This intersects SCAT’s curve on Day 82 (22 March 2020). As mentioned previously, case rates can become unreliable as testing and reporting becomes erratic in a crisis. Instead, looking at death rates can be useful as they can be extrapolated backwards. By 26 March 2020 (Day 86), Italy had reached a case fatality rate of 10%. This may suggest that there is a correlation between critical care bed occupancy and case fatality rate, as the typical survival time of non-survivors is 1–2 weeks. This also intersects SCAT’s curve on Day 82 (22 March 2020), showing a strong similarity to the previously approximated capacity. Figure 11 shows these lines on the case graphs.

**Figure 11:**
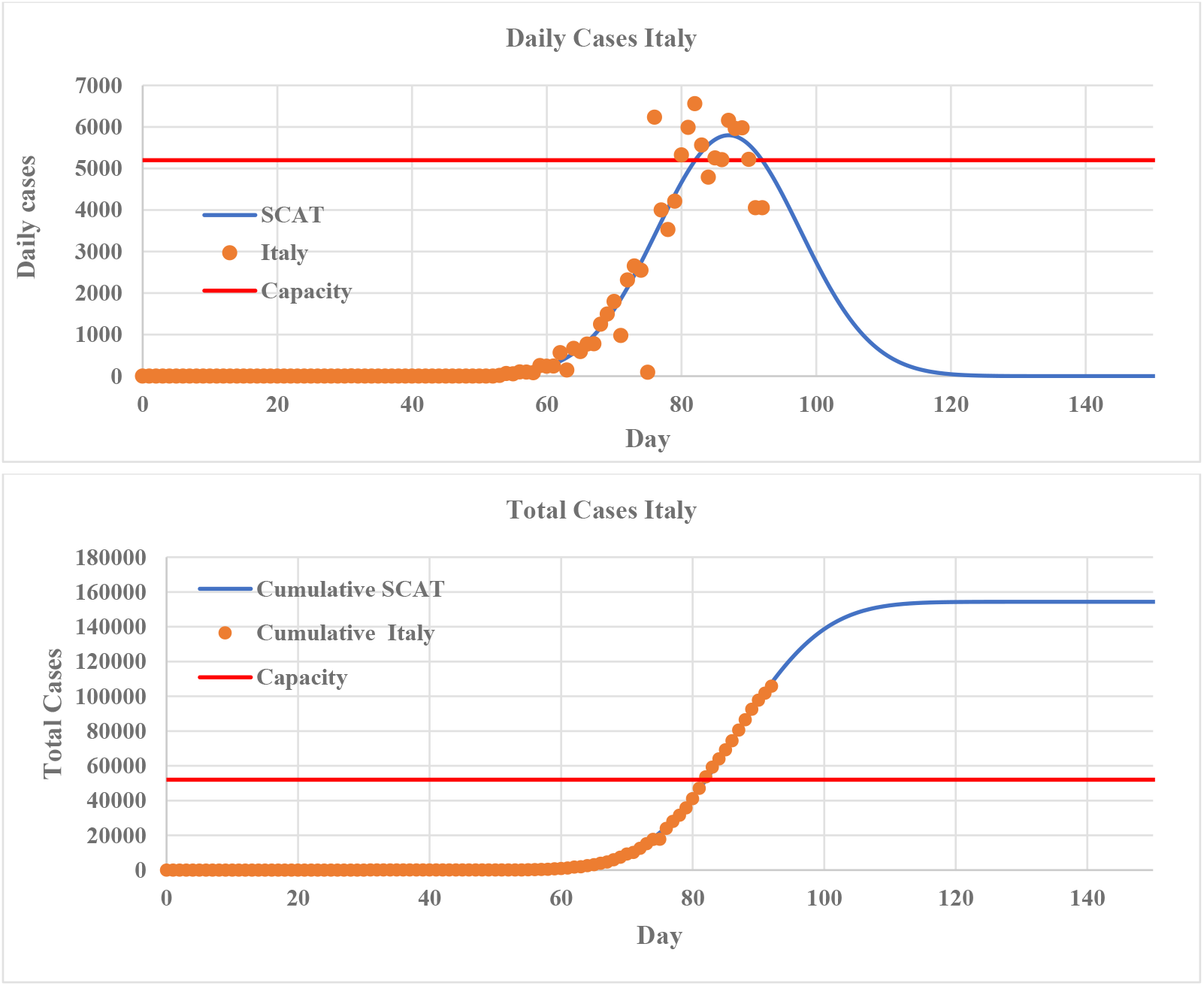
*Approximate capacity lines for Italy on (a) the daily cases curve and (b) the cumulative cases curve, showing normal capacity (before construction of COVID-19 hospitals) reached on approximately Day 82 (22 March 2020).*

In comparison, the UK has fewer hospital beds per capita than most countries in Europe with approximately 101,000 acute care beds and 5,900 critical care beds^24^ which are currently occupied at a rate of 90 %. This leaves just 590 available critical care beds.

### Comparison with Other Diseases

One key confusion in the reporting of this pandemic was the difficulty in understanding the difference between COVID-19 and other similar diseases such as influenza. SCAT allows a clear comparison to be made. In Figure 13, data is taken from the influenza season of 2017-18 in New York^25^ (7 October 2017 to 19 May 2018) which was one of the worst recorded.

**Figure 12:**
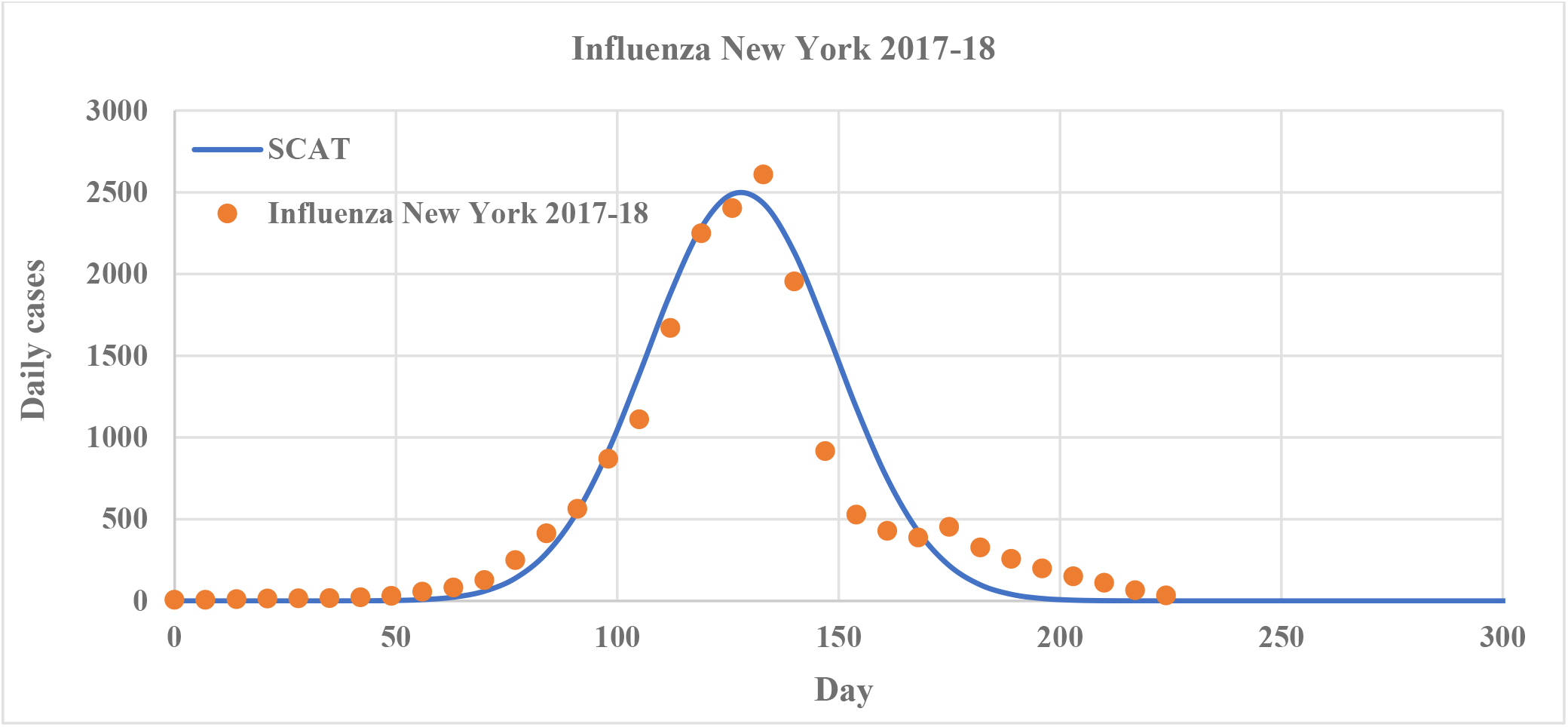
*SCAT applied to data from the influenza season in New York, Oct 2017 to May 2018* (*M* = 2,500, *p* = 128, *FWHM* = 50). *Day 1 = 7 October 2017.*

**Figure 13:**
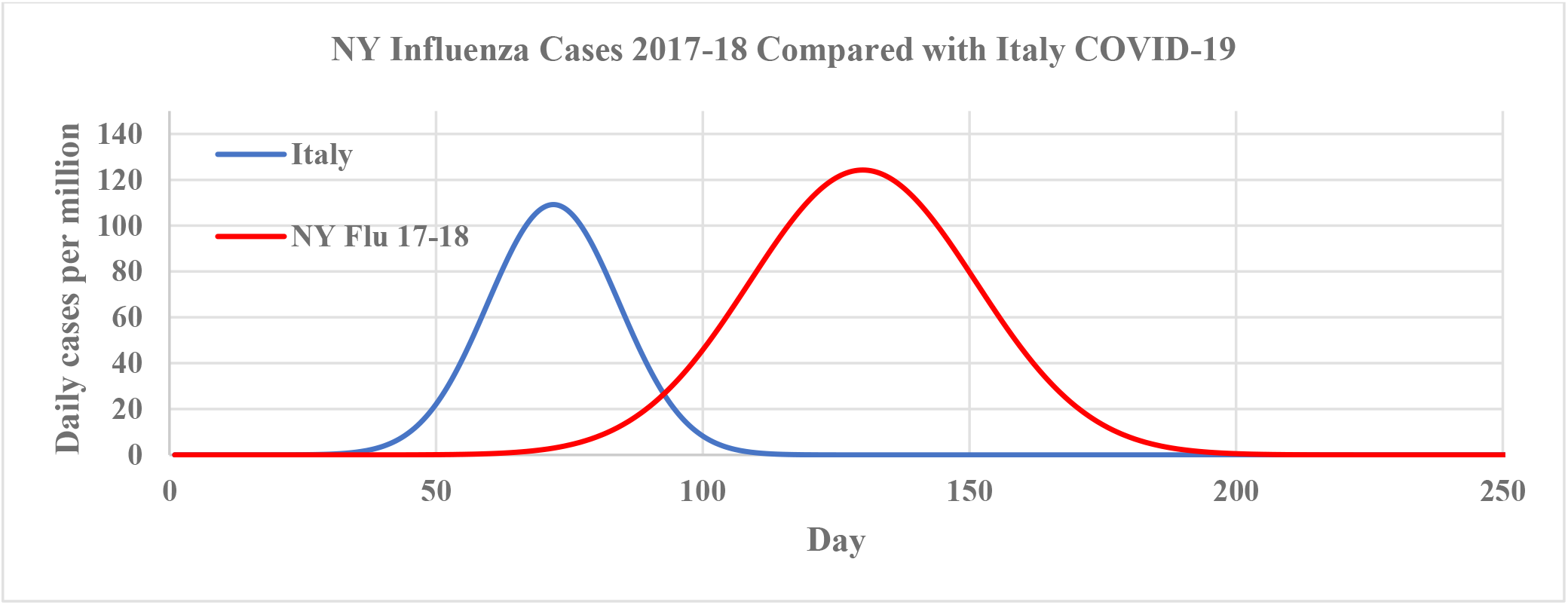
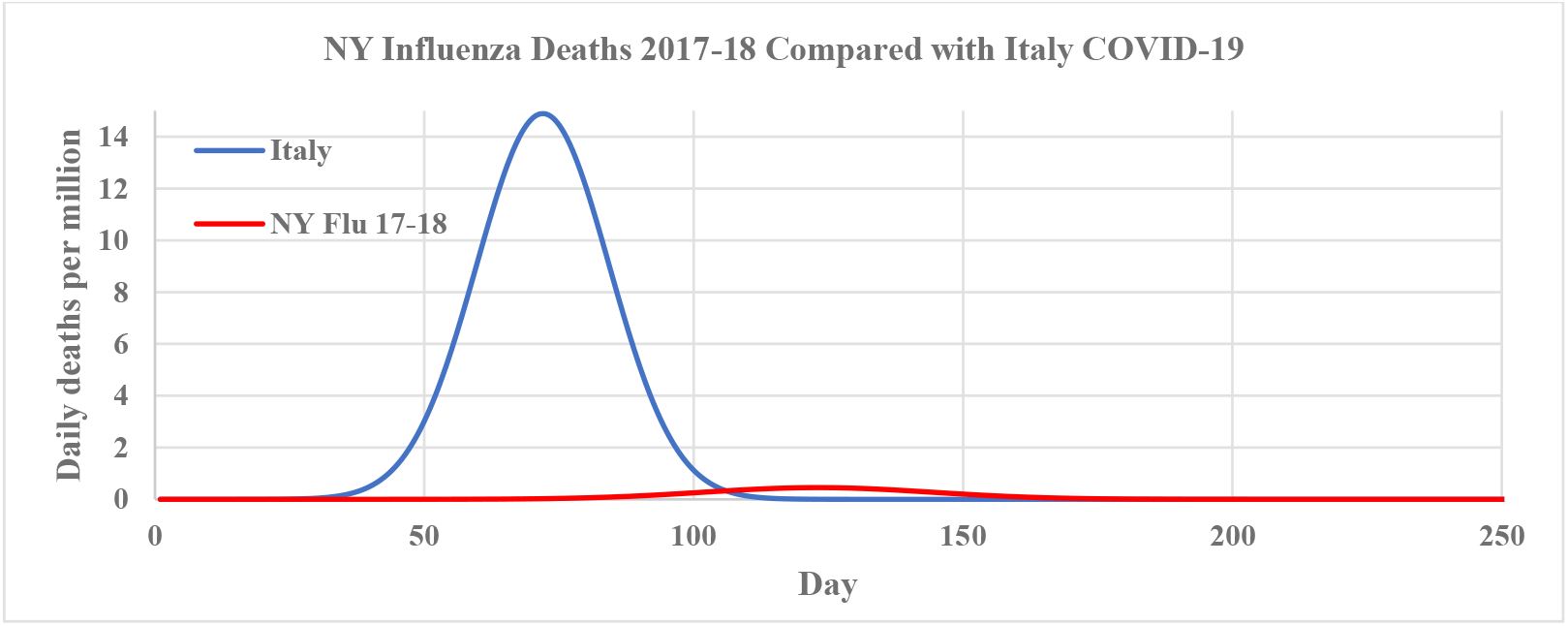
*The curve for the influenza season in New York, Oct 2017 to May 2018 compared with the Italian COVID-19 curve (a) cases (b) deaths per million. The faster speed of COVID-19 spread can be seen in the cases curve but the difference between COVID-19 and Influenza can be most clearly observed in the deaths.*

This curve can now be compared with typical COVID-19 curves. Figure 14 shows the Italian curves compared to the New York influenza curves. At first glance at the case rate, it is understandable why many people see influenza as similar to COVID-19, as influenza appears to have a similar curve and even reaches higher average daily case rates per million (although no attempt is made for the most part to derive causation from results in this work, it should be noted here that strict quarantine measures are in place in Italy while New York was not equivalently locked down during the flu season of 2017-18). However, even here, the faster speed of infection of COVID-19 can still clearly be seen as the peak is reached much quicker than with influenza. But it is in the death rate curve that the stark difference is clearly observed.

**Figure 14:**
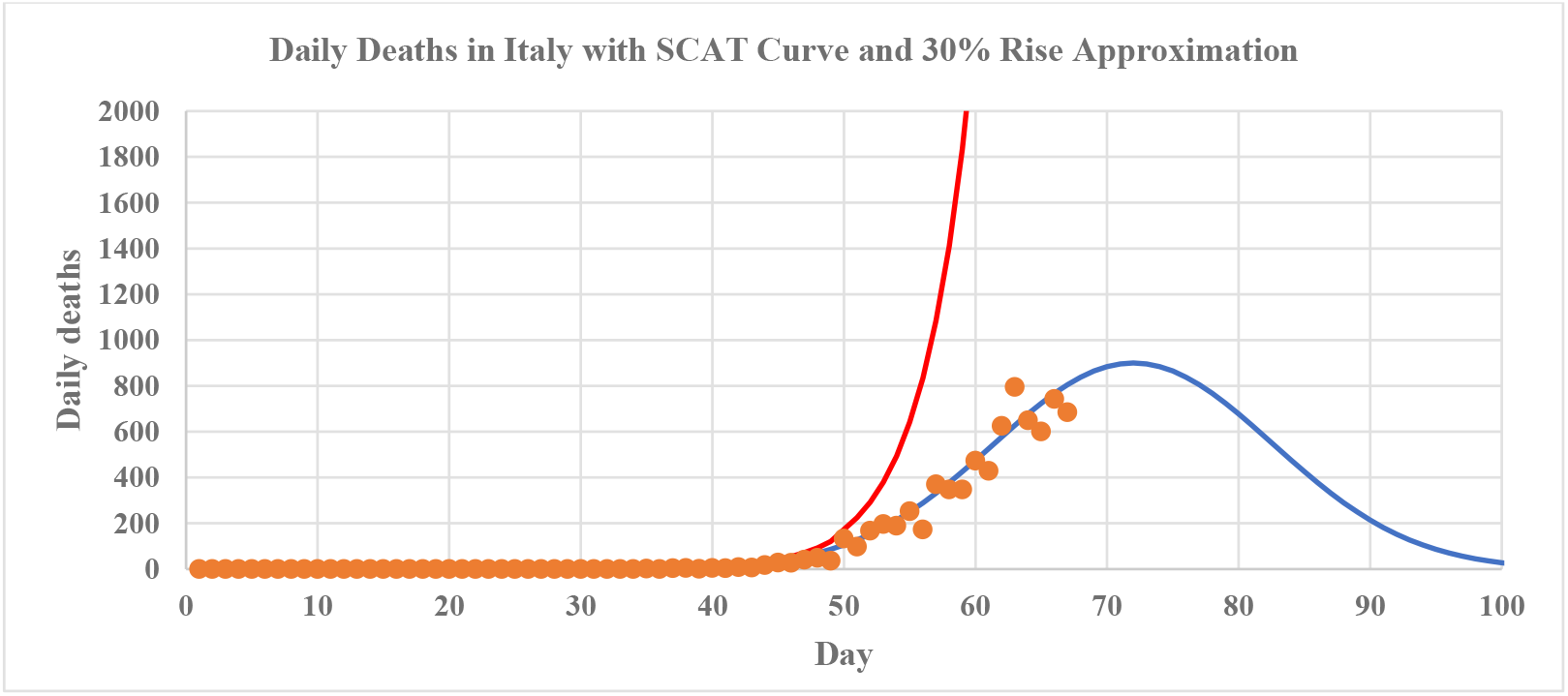
*SCAT applied to Italian death rates compared with the 30% exponential rise used in much of the media reporting.*

A key intention of SCAT is to discredit this kind of misinformation.

## Discussion

Although not a replacement for proper epidemiological modelling, SCAT provides authors, journalists and others with a means to approximate the elusive ‘curve’, and use it to visually compare across countries, diseases, etc. Figure 15 shows the SCAT-generated curve compared with the much published exponential rise curve^26^ and is shown to be a clearly better approximation.

**Figure 15:**
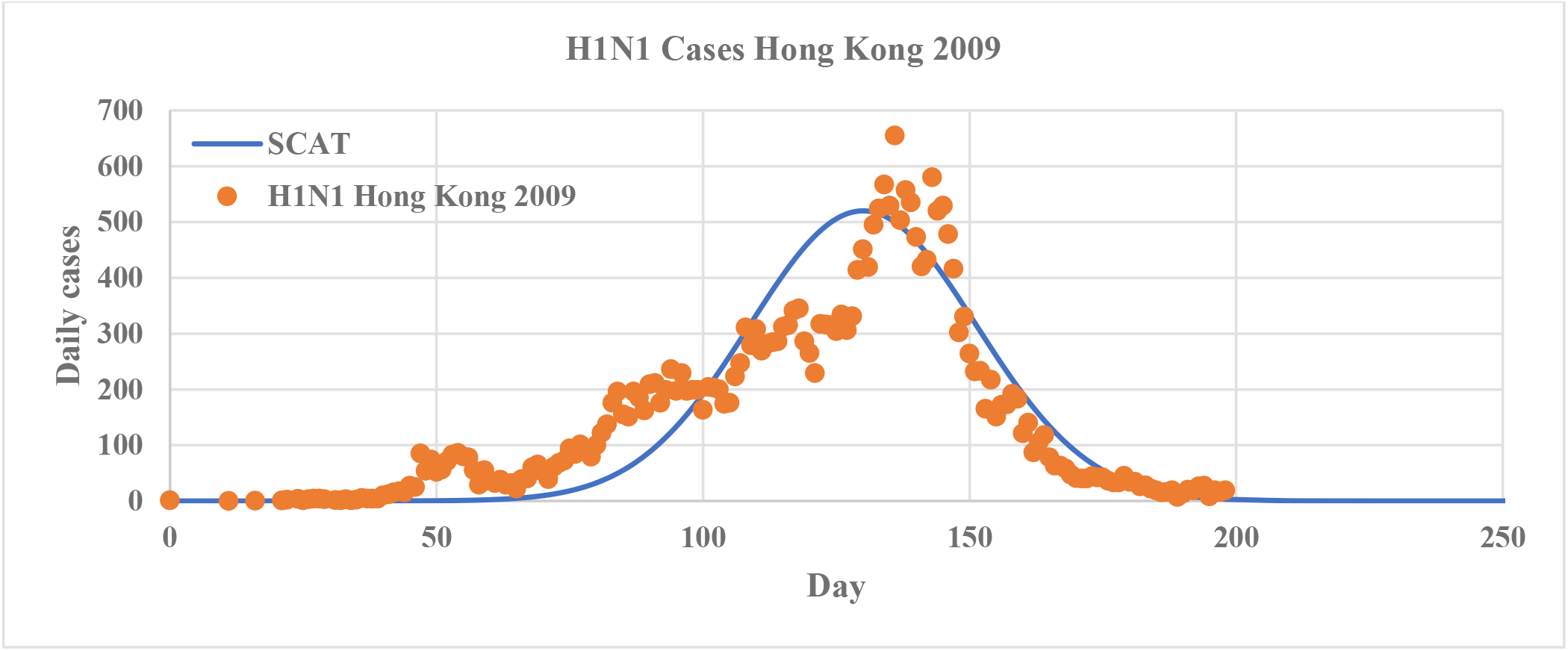
*SCAT applied to the H1N1 epidemic in Hong Kong 2009* (*M* = 520, *p* = 130, *FWHM* = 50). *Day 1 = 1 May 2009.*

### Limitations

SCAT is approximate and is not intended as a replacement for epidemiological models as diseases are rarely well behaved and are likely to stray from the curve. It is also only designed to approximate one ‘season’ of the virus, and not its entire lifetime. Any long-term prediction would be untrustworthy, and variations are expected as mitigation measures are put in place.

To demonstrate this, SCAT was applied to the H1N1 epidemic in Hong Kong^27^ from 1 May 2009 to 15 November 2009. Although SCAT reaches a similar final tally (with less than 0.6 % error), it does not fit the data well in the rise phase as the rise phase is much slower for this disease (a key difference between COVID-19 and other similar pandemics).

The curve does not capture re-infections or seasonal recurrence. Figure 17 shows data for seasonal flu in New York from 2009 to 2019 with SCAT only capturing the 2017-18 season.

**Figure 16:**
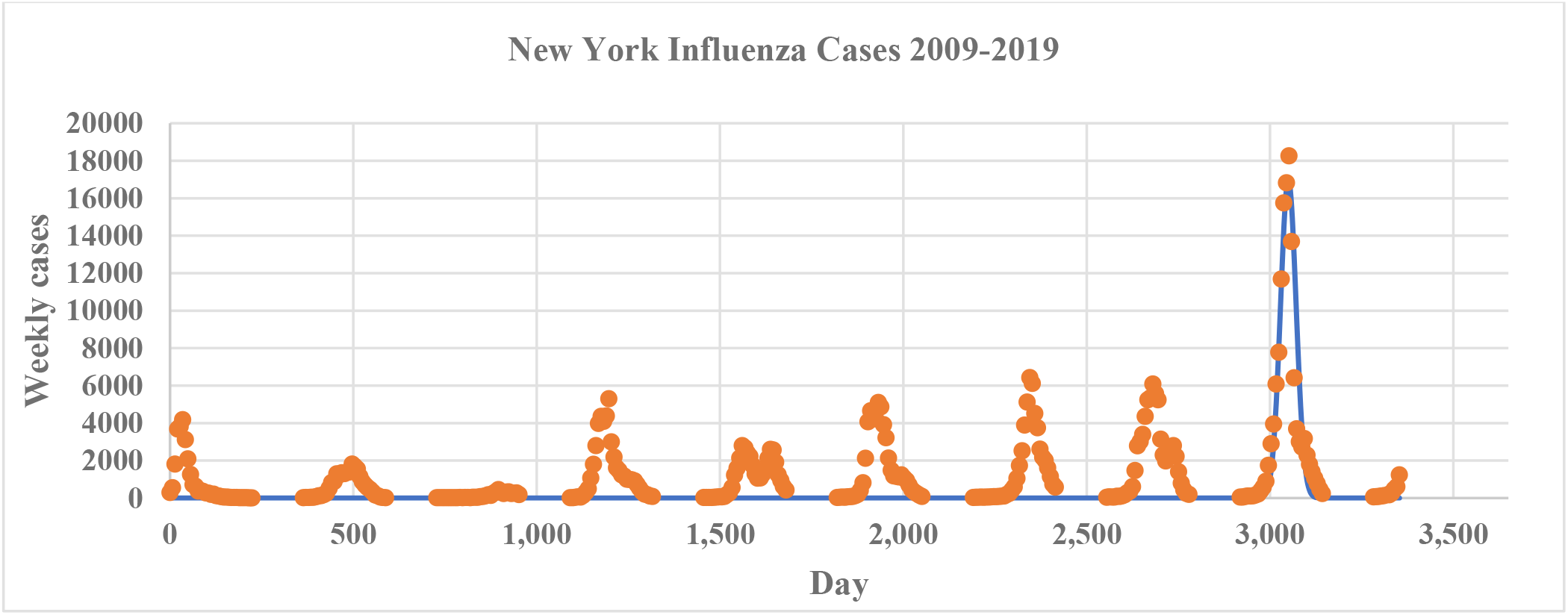
*Seasonal influenza in New York 2009-19 with SCAT capturing only 2017-2018* (*M* = 17,000, *p* = 3050, *FWHM* = 50). *Day 1 = 10 October 2009.*

**Figure 17:**
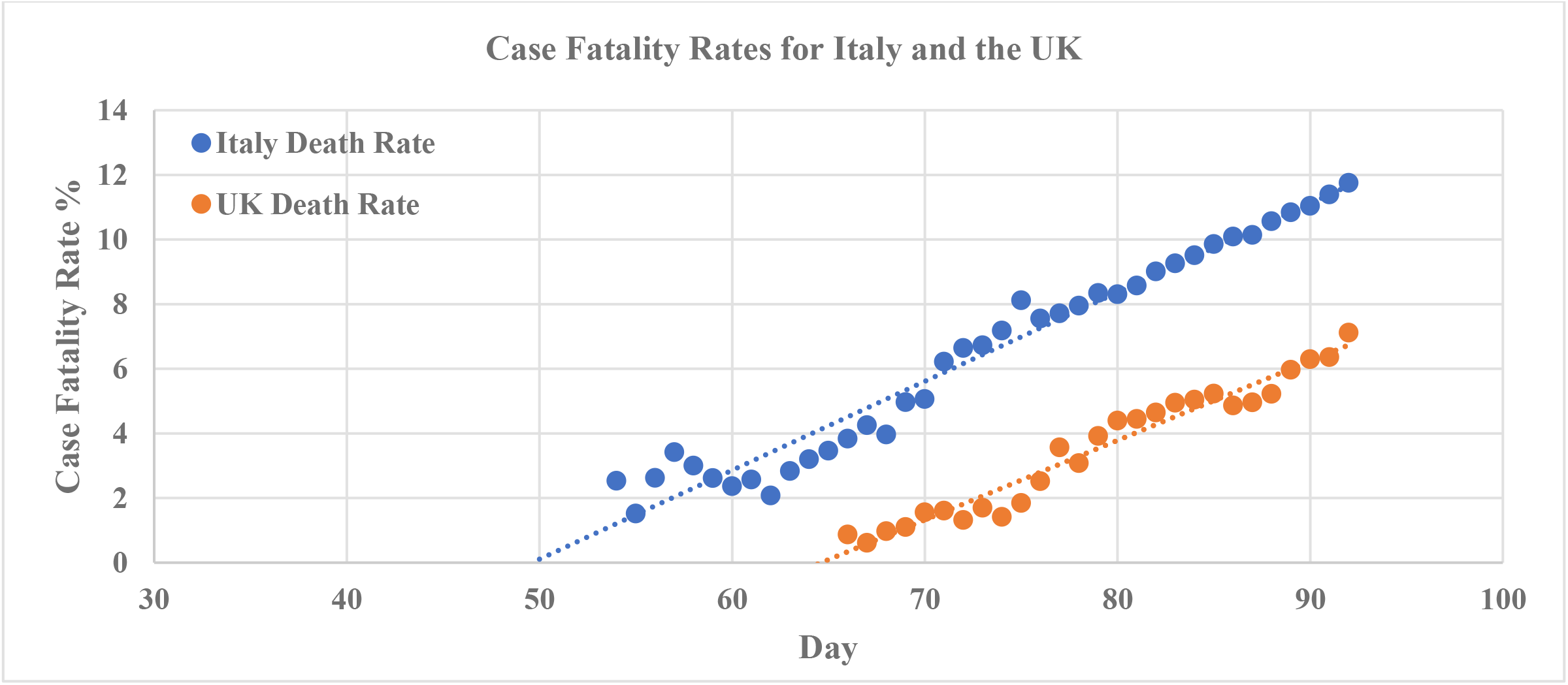
*Rising case fatality rates in Italy and the UK.*

While China’s Case Fatality Rate (CFR) has settled to approximately 4 %, the fatality rates for both Italy and the UK are still rising, with Italy currently at 12 % and UK approaching 8 %, as seen in Figure 12. CFR is not a reliable figure due to inconsistent testing and omission of comorbidity considerations. However, it indicates that there is some way to go before final figures are reached.

### Functionality

SCAT has been shared with epidemiologists, medical editors and lay users and has been found to be very easy to use. Figure 18 shows a snapshot of SCAT in MS Excel. Date and daily cases data are entered in the yellow columns on the left and parameters are entered in the yellow boxes at the top. The rest of the sheet is automatically populated. Guide values for parameters are also automatically generated once the data is entered, to help with parameter estimation.

**Figure 18:**
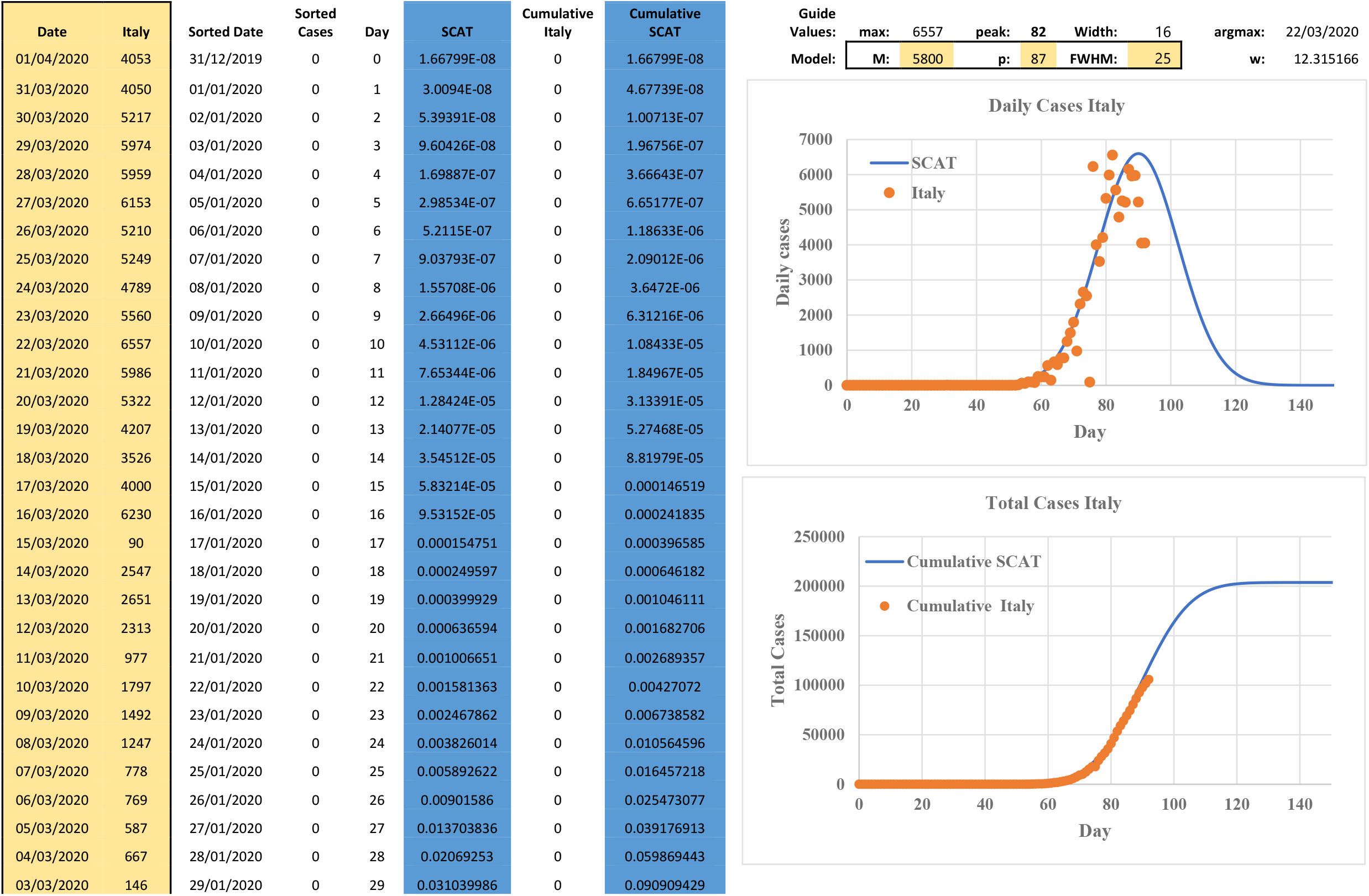
*A snapshot of SCAT in MS Excel.*

It must be reiterated that SCAT is not a replacement for epidemiological modelling and no causation is derived from any observations in this work. It is hoped however, that the ability to approximate the curve will allow faster testing of ideas and clearer dissemination of findings, which will ultimately lead to better understanding of the disease.

## Data Availability

All data is freely available online

## Acknowledgements

I would like to thank Claire Chambers for useful discussions and valuable insights. I hope that these sentences are sufficiently succinct.

## Notes

### Competing Interest Statement

The authors have declared no competing interest.

